# Monitoring drinking water quality in nationally representative household surveys: cross-sectional analysis of 20 Multiple Indicator Cluster Surveys 2014-2019

**DOI:** 10.1101/2020.09.21.20174862

**Authors:** Rob Bain, Rick Johnston, Shane Khan, Attila Hancioglu, Tom Slaymaker

## Abstract

**Background:** The Sustainable Development Goals set an ambitious new benchmark for safely managed drinking water services (SMDW), but many countries lack data on the availability and quality of drinking water.

**Objectives:** To quantify the availability and microbiological quality of drinking water, monitor SMDW and examine risk factors for *E. coli* contamination in 20 low-and middle-income countries.

**Methods:** A new water quality module for household surveys was implemented in Multiple Indicator Cluster Surveys. Teams used portable equipment to measure *E. coli* at the point of collection (PoC, n=48,323) and at the point of use (PoU, n=51,345) and asked respondents about the availability and location of drinking water services. *E. coli* levels were classified into risk categories and SMDW was calculated at the household- and domain-levels. Modified Poisson regression was used to explore risk factors for contamination.

**Results:** *E. coli* was commonly detected at PoC (range 16-90%) and was more likely at PoU (range 20-97%). Coverage of SMDW was 56% points lower than improved drinking water with water quality the limiting factor for SMDW in 14 countries. Detection of *E. coli* at PoC was associated with use of improved water sources (RR=0.64 [0.52-0.78]) located on premises (RR=0.78 [0.67-0.91]) but not with availability (RR=0.94 [0.82-1.06]). Households in the richest quintile (RR=0.67 [0.50-0.90]) and in communities with high (>75%) improved sanitation coverage (RR=0.95 [0.91-0.98]) were less likely to use contaminated water at PoU whereas animal ownership (RR=1.08 [1.03-1.14]) and rural residence (RR=1.11 [1.03-1.19]) increased risk of contamination.

**Discussion:** Water quality data can be reliably collected in household surveys and can be used to assess inequalities in service levels, to track the SDG indicator of SMDW, and to examine risk factors for contamination. There is an urgent need to implement scalable and sustainable interventions to reduce exposure to faecal contamination through drinking water.

## Introduction

The Sustainable Development Goals (SDGs) set ambitious targets for universal access to safe drinking water, sanitation and hygiene (WASH) by 2030. Expert consultations and negotiations between UN Member States led to the adoption of a new global indicator 6.1.1, the "use of safely managed drinking water services” which builds on, but greatly extends, the previous Millennium Development Goal (MDG) indicator of "use of an improved source”. Safely managed drinking water services are defined as improved sources of drinking water (piped water, protected groundwater sources, rainwater collection, packaged or delivered water), that are accessible on premises and provide water which is available when needed and free from faecal and priority chemical contamination (WHO/UNICEF 2017b).

WHO and UNICEF through the Joint Monitoring Programme (JMP) for Water Supply, Sanitation and Hygiene are mandated to monitor the SDG global indicators for WASH and released baseline estimates in July 2017 (WHO/UNICEF 2017b) and updated estimates in 2019 (WHO/UNICEF 2019b). The JMP reports confirm earlier academic studies estimating that approximately 2 billion people drink water from sources that contain faecal indicator bacteria (Bain et al. 2014a; Onda et al. 2012). While the importance of water quality is increasingly recognized by policymakers, the 2019 report found that only four of the eight SDG regions and 98 of the 193 UN Member States had enough data to report on water quality for target 6.1 (WHO/UNICEF 2019b). There is an urgent need to expand the monitoring of drinking water quality to inform national efforts to achieve safely managed drinking water services by 2030.

Globally the highest priority parameter of drinking water quality to monitor from a health perspective is *E. coli*, WHO’s recommended indicator for faecal contamination of drinking water (WHO 2017). Arsenic and fluoride are priority chemical contaminants due to their respective disease burdens. For the purposes of SDG monitoring "free from contamination” implies that drinking water does not contain *E. coli* (in a 100 mL sample) or elevated levels of arsenic (>10 μg/L) or fluoride (>1.5 mg/L).

In preparation for the SDGs, and in response to the recommendations of expert task forces on monitoring drinking water quality reflecting on the limitations of MDG monitoring and the availability of data on water quality from administrative systems (Bartram et al. 2014), the JMP has supported the piloting and scale-up of water quality testing in nationally representative multi-topic household surveys. Integration of water quality testing in household surveys has several key advantages including the ability to obtain representative data at comparatively low cost and to link it with other information on household characteristics collected in the survey (health, nutrition, education, wealth, ethnicity, etc.). Data from household surveys were critical to MDG WASH monitoring and are expected to continue to be a major data source for SDG monitoring alongside data from administrative sources, especially regulators (WHO/UNICEF 2017a).

In close collaboration with UNICEF-supported Multiple Indicator Cluster Surveys (MICS), a new water quality module was developed as MICS transitioned from the fifth global round of surveys (MICS5, 2013-2017) to the sixth round (MICS6, 2017-current). The water quality module is now available as a standard module offered to countries implementing MICS6. This work builds on earlier studies supported by the JMP, in particular the Rapid Assessment of Drinking water Quality (Aldana 2010; Aliev et al. 2010; Howard et al. 2009; Ince et al. 2010; Tadesse et al. 2010). Final reports published by the implementing agencies are available from the MICS website (mics.unicef.org) and researchers have begun to explore the risk factors for contamination (Kandel et al. 2017; Wardrop et al. 2018) and associations with child health and development (Haque et al. 2017). With the exception of Nigeria, without the water quality data made available from household surveys, none of the 20 countries included in this study would have the nationally representative data necessary for monitoring SMDW.

The objectives of this study were to describe the main features of the new water quality module, and to use the data from the first 20 MICS that included the new module to explore patterns in water quality by country, type of water source and wealth, and compare levels of the MDG and SDG indicators for drinking water. We used multivariable regression to explore household- and community-level risk factors for faecal contamination, including accessibility and availability of drinking water.

## Methods

The water quality module was integrated into multi-topic household surveys supported by the global MICS programme and implemented by national authorities, often the National Statistical Offices. Table 1 lists the countries that integrated water quality testing in Multiple Indicator Cluster Surveys between 2014 and 2019 for which microdata are publicly available. The STROBE statement for observational studies informs our reporting and Figure 1 shows how data from the 20 MICS were used in this study.

**Table 1:** Nationally representative MICS that have integrated water testing, 2014-2019.

| Country<br>(MICS round) | Fieldwork | Households<br>interviewed<br>(response rate) | Clusters | Samples at point<br>of collection<br>(response rate) | Samples at<br>point of use<br>(response rate) | Teams | Implementing agencies | Technical assistance for water quality<br>testing | Training<br>duration |
| --- | --- | --- | --- | --- | --- | --- | --- | --- | --- |
| **Bangladesh**<br>(MICS6) | Jan-June<br>2019 | 61242<br>(99.4%) | 3220 | 6069<br>(98.7%) | 6140<br>(99.8%) | 34 | Bangladesh Bureau of Statistics | International Centre for Diarrhoeal<br>Disease Research, Bangladesh | 5 days |
| **Congo**<br>(MICS5) | Nov 2014-Feb 2015 | 12811<br>(99.6%) | 531 | 1277<br>(81.9%) | 1507<br>(95.8%) | 19 | Institut National de la Statistique | Société nationale de distribution d'eau | 5 days |
| **Côte d'Ivoire**<br>(MICS5) | April-July<br>2016 | 11879<br>(96.6%) | 511 | 1782<br>(92.5%) | 1908<br>(99.1%) | 21 | Institut National de la Statistique | Institut National d'Hygiène Publique and<br>Laboratoire National de Santé Publique | 5 days |
| **Gambia**<br>(MICS6) | Jan-April<br>2018 | 7405<br>(98.5%) | 390 | 1764<br>(93.9%) | 1865<br>(99.3%) | 8 | Gambia Bureau of Statistics | Ministry of Water Resources | 5 days |
| **Georgia**<br>(MICS6) | Sept-Dec<br>2018 | 12270<br>(94.2%) | 706 | 2429<br>(79.4%) | 2699<br>(88.2%) | 13 | National Statistics<br>Office of Georgia | National Center for Disease Control and<br>Public Health | 3 days |
| **Ghana**<br>(MICS6) | Oct 2017 –<br>Jan 2018 | 12886<br>(99.4%) | 660 | 3161<br>(98.1%) | 3219<br>(99.9%) | 25 | Ghana Statistical Service | Water Research Institute and Ghana<br>Water Company | 4 days |
| **Iraq**<br>(MICS6) | Mar-May<br>2018 | 20214<br>(99.5%) | 1710 | 6687<br>(99.3%) | 6724<br>(99.9%) | 39 | Central Statistical Organization &<br>Kurdistan Region Statistical Office | Central Public Health Laboratory | 2 days |
| **Kiribati**<br>(MICS6) | Nov 2018 –<br>Jan 2019 | 3071<br>(98.7%) | 164 | 589<br>(94.1%) | 622<br>(99.4%) | 9 | Kiribati National Statistics Office | Ministry of Health and Ministry of<br>Infrastructure and Sustainable Energy | 4 days |
| **Lao PDR**<br>(MICS6) | July-Nov<br>2017 | 22287<br>(99.3%) | 1165 | 3292<br>(98.0%) | 3346<br>(99.6%) | 25 | Lao Statistics Bureau | Center for Environmental Health and<br>Water Supply (Nam Saat). | 4 days |
| **Lesotho**<br>(MICS6) | April-Sept<br>2018 | 8847<br>(95.9%) | 400 | 1339<br>(97.3%) | 1373<br>(99.8%) | 15 | Bureau of Statistics | Ministry of Health and Ministry of Water<br>Affairs | 3 days |
| **Madagascar**<br>(MICS6) | Aug-Nov<br>2018 | 17870<br>(97.7%) | 774 | 3265<br>(94.9%) | 3433<br>(99.8%) | 30 | L'Institut National de la Statistique de<br>Madagascar | Ministre de Santé and the Ministre de<br>l'Eau | 3 days |
| **Mongolia**<br>(MICS6) | Sep-Dec<br>2018 | 13798<br>(98.3%) | 580 | 2598<br>(94.0%) | 2736<br>(99.0%) | 19 | National Statistical Office | UNICEF Mongolia | 3 days |
| **Nepal**<br>(MICS5) | Feb-June<br>2014 | 12405<br>(98.5%) | 519 | 474<br>(96.1%) | 1492<br>(100.0%) | 15 | Central Bureau of Statistics | Environment and Public Health<br>Organization | 7 days |
| **Nigeria**<br>(MICS5) | Sept 2016<br>– Jan 2017 | 33901<br>(98.9%) | 2239 | 2722<br>(89.0%) | 3053<br>(99.8%) | 78 | National Bureau of Statistics | Federal Ministry of Water Resources | 5 days |
| **Paraguay**<br>(MICS5) | June– Sept<br>2016 | 7313<br>(96.3%) | 499 | 1750<br>(96.5%) | 1790<br>(98.7%) | 15 | Dirección General de Estadística,<br>Encuestas y Censos | Ente Regulador de Servicios Sanitarios | 2 days |
| **Sierra Leone**<br>(MICS6) | May-Aug<br>2017 | 15309<br>(99.6%) | 600 | 1748<br>(98.0%) | 1780<br>(99.8%) | 24 | Statistics Sierra Leone | Ministry of Water Resources | 5 days |
| **Suriname**<br>(MICS6) | Mar-Aug<br>2018 | 7915<br>(90.2%) | 470 | 1619<br>(81.7%) | 1701<br>(85.8%) | 10 | General Bureau of Statistics | Suriname Water Supply Company and the<br>Bureau of Public Health | 4 days |
| **Togo**<br>(MICS6) | July-Oct<br>2017 | 7916<br>(98.2%) | 420 | 1088<br>(94.0%) | 1153<br>(99.7%) | 16 | Institut National de la Statistique et<br>des Etudes Economiques et | Institut National d'Hygiène | 5 days |
|  |  |  |  |  |  |  | Démographiques |  |  |
| **Tunisia**<br>(MICS6) | Mar-May<br>2018 | 11225<br>(97.8%) | 600 | 2664<br>(95.0%) | 2769<br>(98.7%) | 32 | Institut National de la Statistique | Ministry of Health, Department of<br>Environmental Hygiene and<br>Environmental Protection | 3 days |
| **Zimbabwe**<br>(MICS6) | Jan-Apr<br>2019 | 11091<br>(98%) | 462 | 2043<br>(95.6%) | 2124<br>(99.3%) | 17 | Zimbabwe National Statistics Agency | Ministry of Health and Child Care | 4 days |
| **TOTAL** | - | 311655<br>(98.3%) | 16620 | 48360<br>(94.2%) | 51434<br>(98.3%) | 464 | - | - | - |

**Figure 1.**
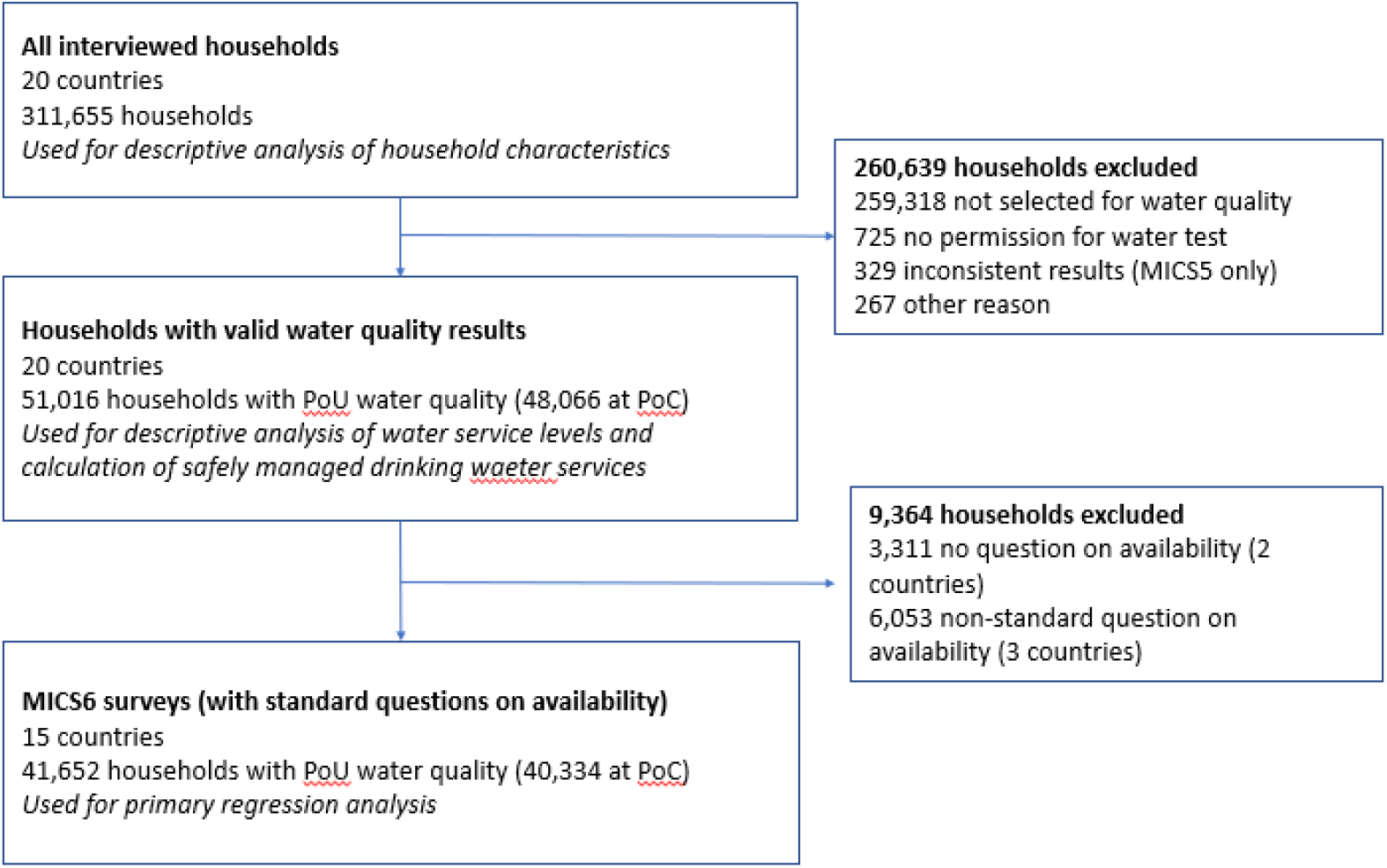
Study flow diagram.

### Water quality module

The water quality module was developed as a collaboration between the JMP and the global MICS programme and was first piloted in Bogra and Sirajgani districts, Bangladesh in April/May 2012 prior to being adopted in the national MICS Bangladesh survey in 2012-2013. The procedures have evolved since the pilot study and further details are provided in the respective survey final reports. The procedures for the standardized module are described in the MICS Manual for Water Quality and MICS Water Quality Testing Questionnaire, both available on the MICS website (mics.unicef.org/tools).

In households selected for water quality testing, the interviewer requests permission to collect samples of drinking water, asking the respondent for a *"glass of drinking water’* (point of use – PoU) and to be shown the location of the source of the drinking water (point of collection – PoC) to take a sample from that location. Two samples are analyzed to examine differences in quality between the point of collection (PoC) and the point of use (PoU). The water quality testing questionnaire includes sections for the interviewer to record test results. It also includes specific questions on the source of the glass of drinking water and household water treatment and an observation of water storage practices.

The field team member responsible for water quality testing differs between surveys depending on the distribution of other tasks. In MICS, the "measurer” who is responsible for anthropometry usually also conducts the water quality test, but in some countries the male interviewer or supervisor were selected instead.

### Water quality test procedures

Water samples were analyzed for *E. coli* using a portable membrane filtration apparatus (Figure S1). *E. coli* was measured by field teams, who filtered 100 mL of sample through a 0.45 micron filter (Millipore Microfil®) which was then placed onto CompactDry™ EC growth media plates (Nissui, Japan) which had been rehydrated with 1 mL of sample water. In the first six surveys (MICS5), an additional 1 mL sample was also tested directly onto a second media plate. Water samples were either collected directly into the pre-assembled manifold or using sterile Whirl-Pak sample collection bags (Enasco). Samples were analysed on-site within 30 minutes and incubated overnight (24-48 hours) in either an electric incubator (Lynd) or incubation belt or pouch specifically designed to keep samples at close to body temperature. The number of *E. coli* (blue) and total coliform (red/purple) colonies were enumerated by eye the following day and recorded in the water quality testing questionnaire. Survey teams recorded cases where there were more than 100 colonies on a given plate and in cases where the plate turned pink as ‘too numerous to count’. For more recent surveys (MICS6), *“E. coli* only” CompactDry™ plates were used, with the advantage that the second media plate was not needed to count up to 100 *E. coli*.

### Sample design for the water quality module

MICS are based on a multistage stratified cluster survey design in which households have a known probability of selection enabling national estimates to be generated (Khan and Hancioglu 2019). To ensure an adequate sample size for sub-national regions, the sample is usually stratified (for example urban/rural and geographic regions). MICS usually include informal urban settlements or slums in sampling frames, but are rarely designed to stratify results for these settings. Within each stratum, enumeration areas are randomly selected from which 20-25 households are selected for inclusion in the survey. Enumeration areas vary between countries but are often between 100 and 250 households. The water quality module was conducted in a sub-sample of households (between 1 and 5) within each cluster. Not every household was selected to minimize the cost and workload and because it was anticipated that the marginal gains of increasing the sample size would be small due to intra-cluster correlation (households in the same cluster having similar water quality) and correspondingly high design effects. It was anticipated that this would be particularly the case for households using the same community water source (Giné-Garriga et al. 2013). Design effects were calculated and are reported in supplementary materials (Table S1).

### Training and QA/QC

An international water quality trainer working alongside national experts led training of field teams. A standard training programme was used and adapted depending on the size of the survey and number of field teams. For example in Nigeria a centralized training of trainers was followed by zonal trainings by microbiologists selected by the Federal Ministry of Water Resources (FMWR). A hands-on training course lasting between 2 and 7 days usually included field practice, a strong focus on aseptic procedure and understanding the different scenarios that testers might face through role-play. National experts from regulatory agencies, research laboratories or ministries of health provided technical inputs to the training and oversight during the fieldwork with a small number of visits scheduled to visit field teams to allow for observation of the testing procedure.

A key measure of quality control used in water quality analysis is a "blank test”, a sample utilized to ensure that the analyst has not inadvertently contaminated the sample and is able to consistently produce the expected negative results. In each survey blank tests (usually one per cluster) were conducted by the field teams to provide confidence in the results. Water for the blank test was obtained from a reliable brand of mineral water or distilled/deionized water. Blank tests using bottled mineral water or deionized water from a laboratory were used as a negative control in the surveys (Table S2). The proportion of positive blanks was ≤2.5% in 18 out of 20 countries indicating high levels of compliance with testing procedures, but there were an elevated number in Gambia (6.2%) and Cote d’Ivoire (8.2%). Two blank tests in Cote d’Ivoire and four in Gambia exceeded 10 *E. coli* per 100 mL.

### Household questionnaire

In addition to developing the new water quality module, the household questionnaire (available from mics.unicef.org/tools) was adapted to include a new question on the availability of drinking water. In the MICS5 round different questions were asked:

> *Nepal*. Since last (day of week) did you have water coming from the pipe or tap for at least one hour a day? *(piped water only)*
>
> *Nigeria*. Was the water from this source not available for at least one full day? *(excludes piped on premises)*
>
> *Paraguay*. In the last 15 days has there been any time you have not been able to access water in sufficient quantities? (*translation from Spanish*)

The more recent MICS6 surveys have used the following standard question on the availability of drinking water aligned with the JMP core questions (WHO/UNICEF 2018b):

> In the last month, has there been any time when your household did not have sufficient quantities of drinking water?

In all surveys, households were asked where their main drinking water source was located and the roundtrip travel time to collect drinking water. In all countries the main source of drinking water and the source of the water quality sample (where this differed) were recorded (Table S4). In most cases the water sample was taken from the household’s main source of drinking water or the same classification of water source used in this study but there were notable differences in some countries, particularly Kiribati (27%) and Mongolia (16%).

### Descriptive analysis

Microdata were downloaded from the MICS website (mics.unicef.org/surveys). Data analysis was conducted using Stata 16 (Statacorp). For each survey descriptive statistics and 95% confidence intervals (95% CI) were calculated using appropriate sample weights and taking into account the stratification and clustering using the *svy* command. For most surveys, separate sample weights were calculated for the source and household water quality samples due to different sub-samples used or response rates. Information on drinking water quality was combined with data from the household questionnaire (type of water source, household assets) to examine patterns in water quality for different population sub-groups and calculate the proportion of the population using safely managed services.

The levels of *E. coli* were classified according to the number of colony forming units (CFU) per 100 mL as follows: <1 "Low risk”, 1-10 "Moderate risk”, 11-100 "High risk” and >100 "Very high risk” (WHO and UNICEF 2012; WHO 2017). There was some evidence for heaping of the last digit in a few countries (Figures S2-S4) but we deemed this insufficient to require adjustment of the risk classification (Wardrop et al. 2018). In a small number of cases from MICS5 datasets, results of the 100 mL and 1 mL tests were inconsistent, could not be classified according to these categories and data were excluded from the analysis (Table S5).

### Calculation of safely managed drinking water services

Safely managed drinking water services are defined as an improved source of drinking water, accessible on premises, available when needed and free from faecal and priority chemical contamination (United Nations 2017). For the countries with information on accessibility, availability and quality (n=17) we estimated the proportion of the national population using improved drinking water sources (MDG indicator) and safely managed drinking water services (SDG indicator) taking into consideration all three criteria. For the remaining countries (n=3) all improved water sources meeting the quality and accessibility criteria were assumed to be available when needed. We calculated estimates for safely managed services based on information from individual households ("household-level”) and also using the approach taken by the JMP whereby a minimum of the three new criteria for safely managed is used at the domain-level ("domain-level”). For global monitoring the JMP uses the "domain-level” approach to accommodate estimates drawn from different data sources, as relatively few countries have all of the information required to calculate the new safely managed indicator from a single data source and at the household level (WHO/UNICEF 2018a).

### Risk factors for contamination

To explore risk factors for faecal contamination of drinking water at the PoC and PoU we used bivariate and multivariable modified Poisson regression models with results reported as risk ratios (RRs) (Zou 2004). Modified Poisson regression is particularly suited to indicators with a high prevalence and has been used in recent studies of handwashing and the association between water and sanitation and trachoma (Garn et al. 2018; Wolf et al. 2019). The dependent variable in these models was the proportion of the population with drinking water contaminated (≥1 CFU/100 mL) or very highly contaminated (>100 CFU/100 mL) with *E. coli* at the PoC or PoU. Pooled estimates were generated using a multilevel model with random intercepts for each country (Gelman and Hill 2007). Random intercepts for clusters were not included as these had a negligible impact on the pooled effect sizes and their confidence intervals in models that converged. Separate country-level models with cluster random intercepts were fit to examine their contributions to the pooled estimates and calculate unadjusted and adjusted RRs for each country. To visualize the pooled and country-level regression results we plotted coefficients and their 95% CIs.

To examine the relationship between the criteria for safely managed services, the presence of *E. coli* at the PoC and PoU was first predicted by whether the water source was improved, accessible on premises and available when needed (using only MICS6 surveys with standardized questions on availability). In a second set of models, we differentiated types of improved water source (piped, packaged, boreholes/tubewells, delivered water, protected wells and springs) and examined a wider range of pre-defined household-level (residence, wealth quintile, improved sanitation, shared sanitation, open defecation, handwashing, education of household head, sex of household head and animal ownership) and cluster-level (>75% improved sanitation, no open defecation) variables in these models (see Table S5). Additional risk factors for contamination at PoU included: natural flooring, handwashing facilities with water and soap, water storage, and appropriate household water treatment. Variables were excluded from country-level models where there were fewer than 25 cases, for example for open defecation in Georgia or use of boreholes as a main source of drinking water in Suriname and Kiribati.

We conducted sensitivity analyses to assess the robustness of the regression models to alternative specifications including the use of custom wealth index quintiles excluding WASH variables (Martel 2017) and the source of the sample of drinking water rather than the household’s main source.

## Result

### Survey characteristics

The total number of households interviewed ranged from 3,280 (Kirbati) to 64,400 (Bangladesh) with a total sample size of >300,000 households (Table 1). Most selected households with completed household interviews provided a sample of drinking water at PoU (mean 98%, range 86-100%) and at PoC (mean 94%, range 79-100%). The response rate is lower for samples at PoC, often as a result of water sources being too far, not being accessible or not having water available at the time of interview (Table S6). The number of water quality samples collected ranged from 622 to 6,724 at PoU (n=51,434 in total) and 454 to 6,687 at PoC per country (n=48,360 in total).

Tables S7 and S8 summarize the main characteristics of water services and pre-defined risk factors for faecal contamination in the 20 surveys that have integrated water quality testing. The proportion of the population using improved drinking water sources (the MDG indicator for drinking water) ranges from 43% in Madagascar to over 95% in Bangladesh, Georgia, Iraq, Paraguay, Suriname and Tunisia. Drinking water was accessible on premises for >75% of the population in these six countries and in Lao PDR but below 50% in 9 countries. Availability of drinking water when needed ranged from 68% in Kiribati to >95% in Bangladesh and Lao PDR. Households were most likely to report appropriate treatment practices (boiling, filtration or adding chlorine) in Kiribati (88%), Mongolia (83%) and Lao PDR (37%). Where respondents were observed filling a glass of drinking water, this was often from a storage container in countries that recorded this information (>50% in 12 out of 17 countries) but the respondent usually obtained water directly from the water source in Georgia, Iraq, Tunisia and Suriname.

Improved sanitation facilities were used by over half of the population (>75% in 7 countries) except in Madagascar and Togo, where open defecation was practiced by 40% and 45% respectively. The proportion of the population with handwashing facilities with water and soap ranged from 12% to 92% and was <50% in 9 countries. The proportion of household heads having secondary education or higher ranged from 16% in Nepal to 90% in Georgia. Around half of countries are predominantly rural whereas in 5 out of 20 countries over two-thirds of the population lives in urban areas. Animal ownership ranged from 11% in Suriname and 14% in Iraq to 89% in Kiribati and natural flooring ranged from <1% in Georgia and Tunisia to 61% in Bangladesh and 67% in Nepal.

### Water quality results at national level

Figure 2 shows the risk levels for *E. coli* in the source of drinking water (point of collection). The proportion of the population using a drinking water source (either improved or unimproved) with detectable *E. coli* (>=1 *E*. coli per 100 mL) ranged from to 16% [14-19] in Mongolia and 20 [18-23] in Tunisia to 85% [80-89] in Kiribati and 90% [88-93] in Sierra Leone. A substantial proportion of the population used very high risk drinking water sources (>100 *E. coli* per100 mL) in Madagascar (51% [48-54]), Sierra Leone (49% [45-53]), Nigeria (46% [43-49]), Togo (38% [33-42]), Lao PDR (36% [34-38]), Kiribati (35% [29-40]) and Côte d’Ivoire (34% [30-38]).

**Figure 2:**
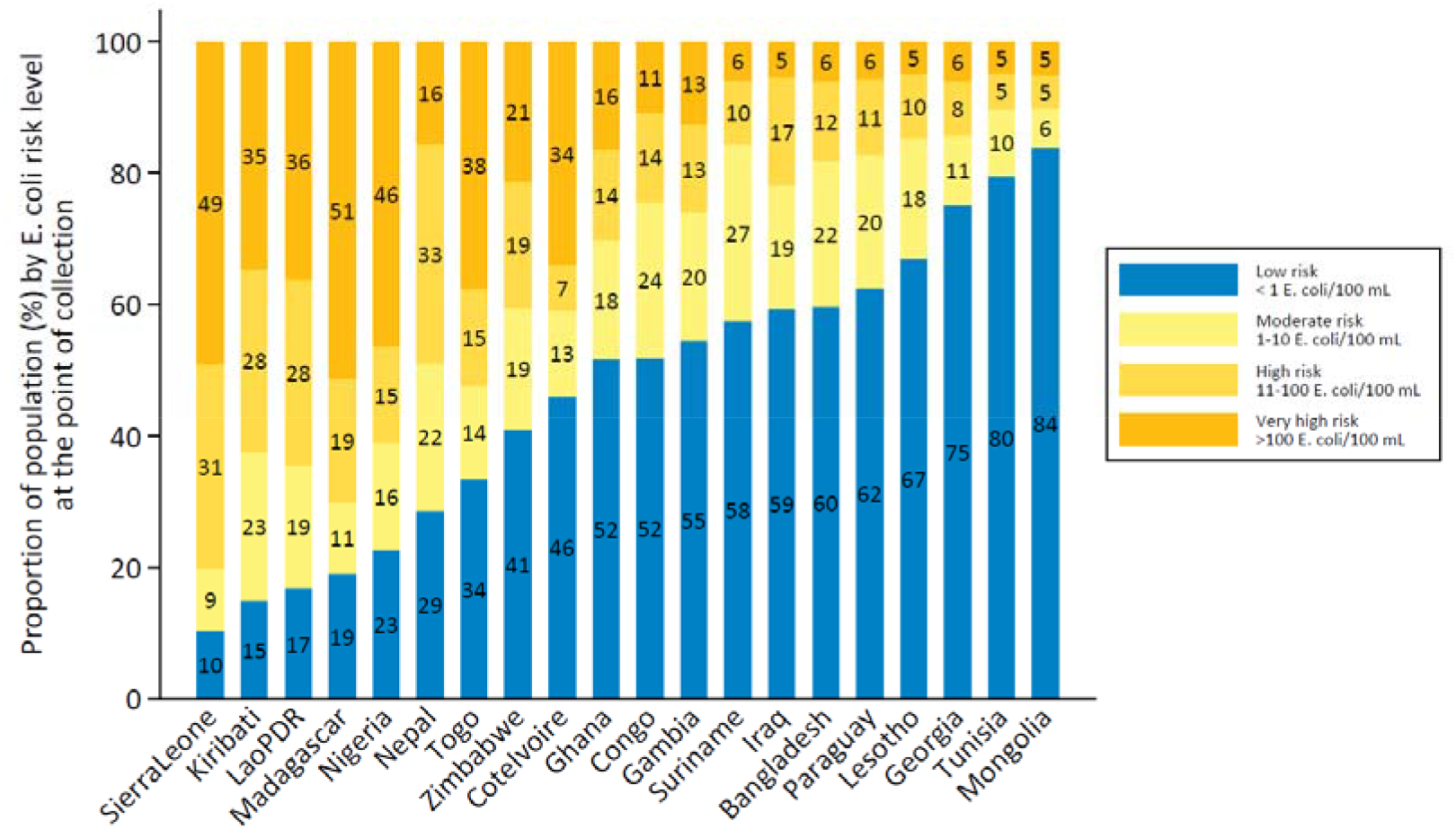
Proportion of population by *E. coli* risk level at the point of collection.

Figure 3 shows the risk levels for *E. coli* in the glass of drinking water within the home. The proportion of the population consuming drinking water with detectable *E. coli* at PoU ranged from 20% [17-23] in Mongolia to 97% [96-98] in Sierra Leone and over half the population were exposed to very high risk drinking water at PoU in Madagascar, Nigeria, Sierra Leone and Togo. In all countries drinking water was more likely to be contaminated at the PoU than PoC.

**Figure 3:**
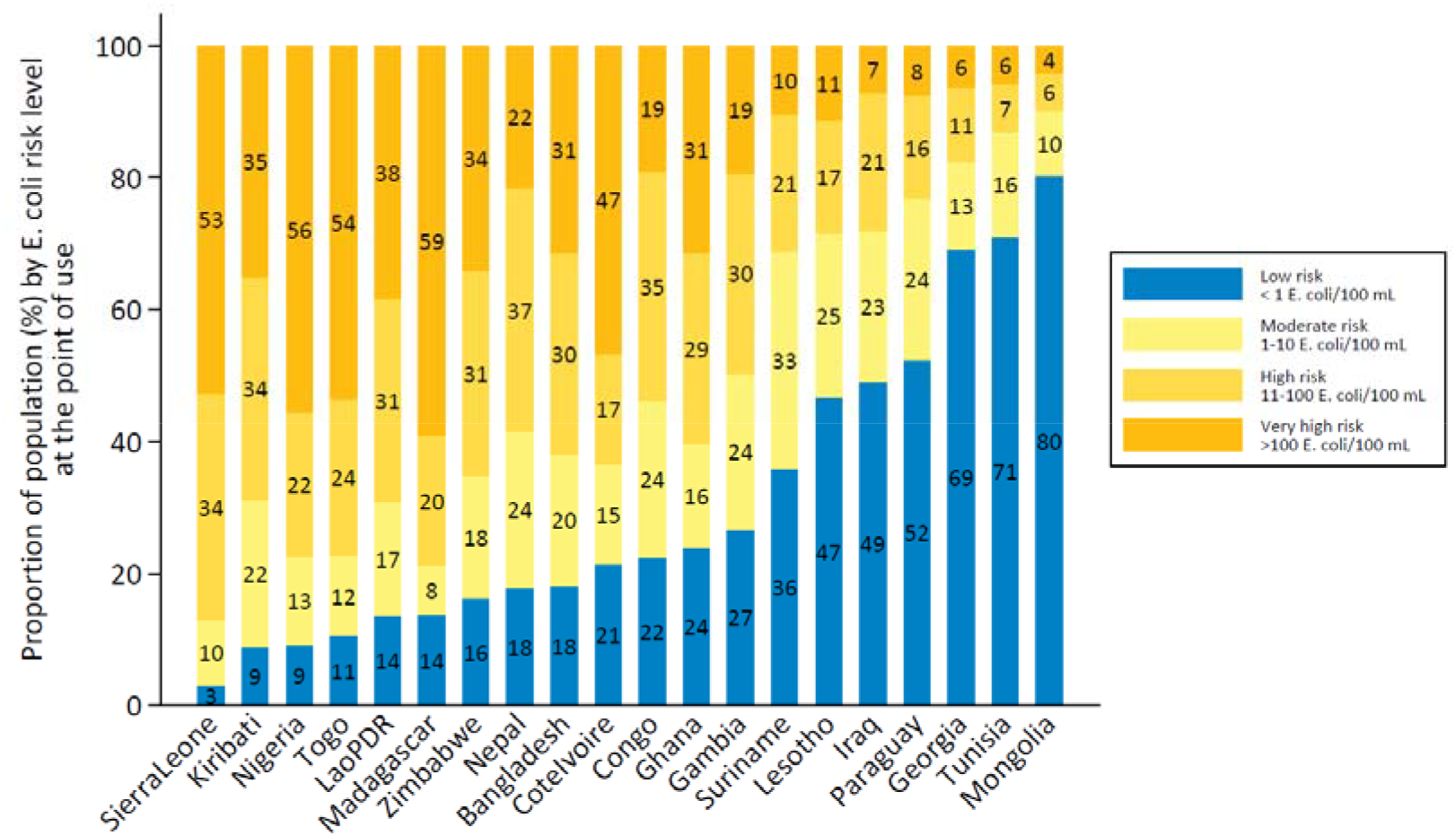
Proportion of population by *E. coli* risk level at the point of use.

### Change in water quality between point of collection and use

For the households with paired samples at PoC and PoU, it is possible to assess whether water quality deteriorated, stayed the same or improved between collection and use (Figure S6). A similar pattern is seen across the countries: deterioration (10-66%) in water quality was more likely than improvement (3-18%) although most often the risk level did not change (30-87%). Among households using a contaminated drinking water source and reporting appropriate water treatment practices, water quality improved in less than half cases (13-44%), except in Mongolia where water quality often improved (70%). As a result, fewer than one in five of these households used water free from *E. coli* at PoU in 17 out of 20 countries and more than one in five used very high risk drinking water at PoU in 14 out of 20 countries (Figure S7).

### Water quality, accessibility and availability by water source type

Figure 4 shows that the levels of *E. coli* vary considerably between water source types and between countries, but that contamination in all source types ranged from low to very high. The general pattern suggests that piped water was often the most likely be free from contamination, followed by water from boreholes/tubewells then packaged and delivered water. The quality of piped water varies considerably between countries with around 20% of the population using piped water that exceeds 100 *E. coli* per 100 mL in Bangladesh, Kiribati, Nepal and Nigeria, and around 40% in Sierra Leone and Lao PDR. In Bangladesh, boreholes provided considerably higher quality water than piped water supplies (63% vs 44% free from *E. coli)*. Other improved sources were also often found to be contaminated. Rainwater and protected wells and springs consistently had significant levels of contamination but appear to be less frequently contaminated than unimproved sources such as unprotected wells and springs and surface water. Delivered water in Mongolia and Tunisia was usually free from *E. coli* but this was not the case in Iraq and Nigeria. Figure S8 shows the *E. coli* risk levels at PoU.

**Figure 4:**
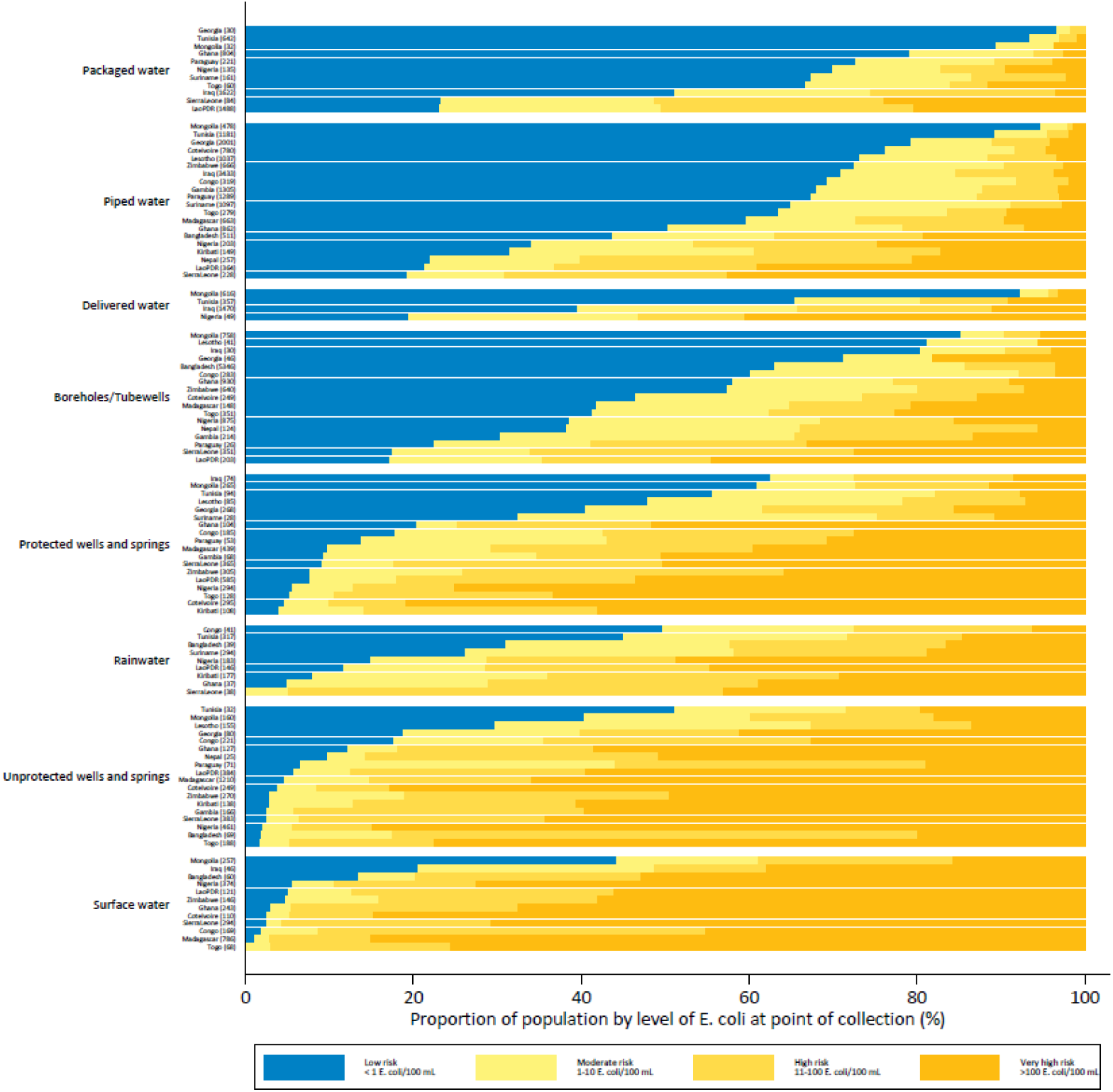
*E. coli* at the point of collection, by type of drinking water source. *Note: For source types with at least 25 cases. Water source categories ordered by unweighted mean proportion with no detectable E. coli*.

Figure 5 shows that there is also considerable variation in accessibility of different water source types across countries, but unimproved sources and surface water are less likely to be accessible on premises or within 30 minutes.

**Figure 5:**
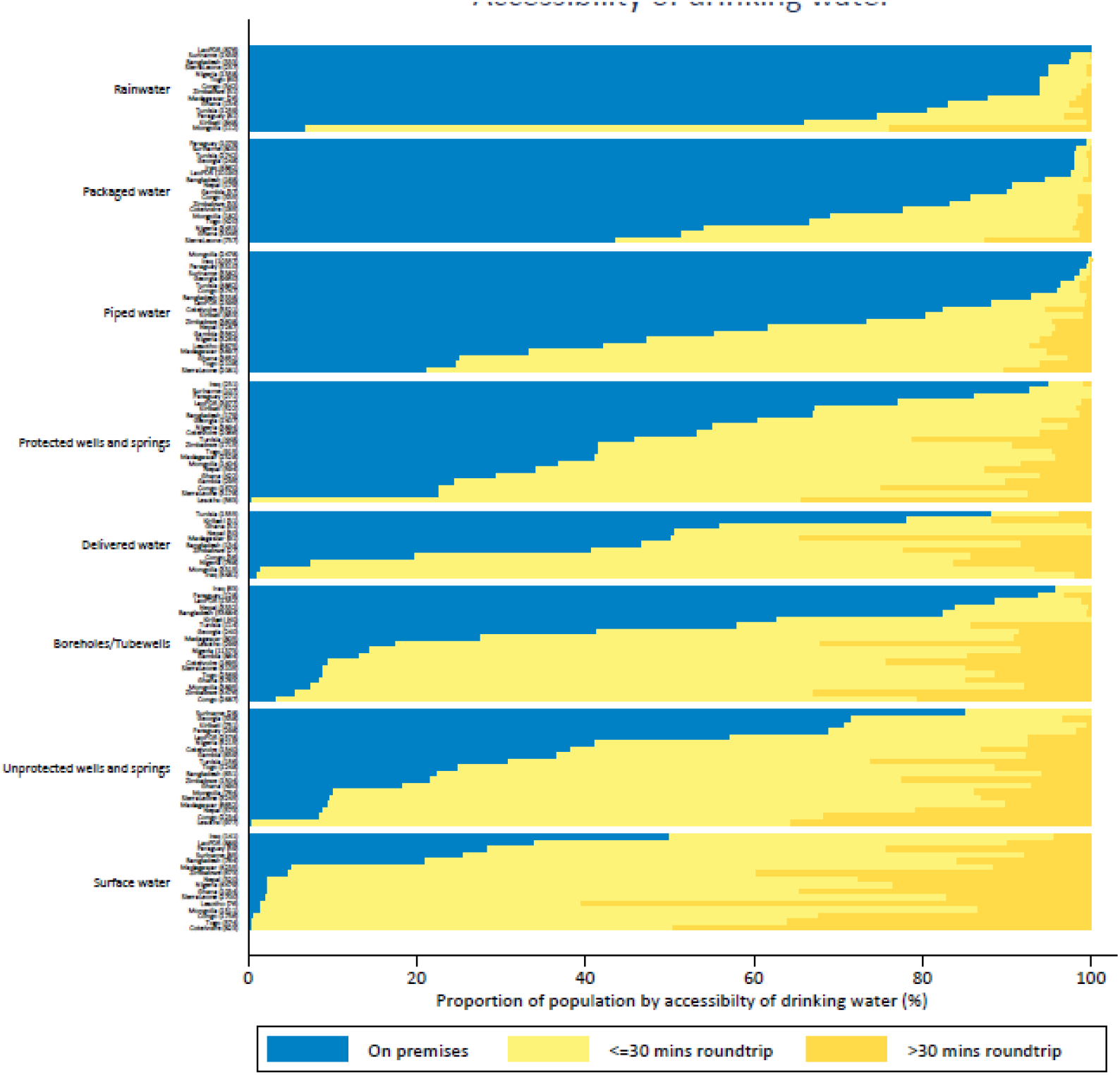
Location and time to collect water from of main source of drinking water, by type of water source. *Note: For source types with at least 25 cases. Water source categories ordered by unweighted mean proportion with on premises*.

Figure 6 shows that there is less variability in reported availability of drinking water, but piped water is often less reliable than other improved and unimproved sources.

**Figure 6:**
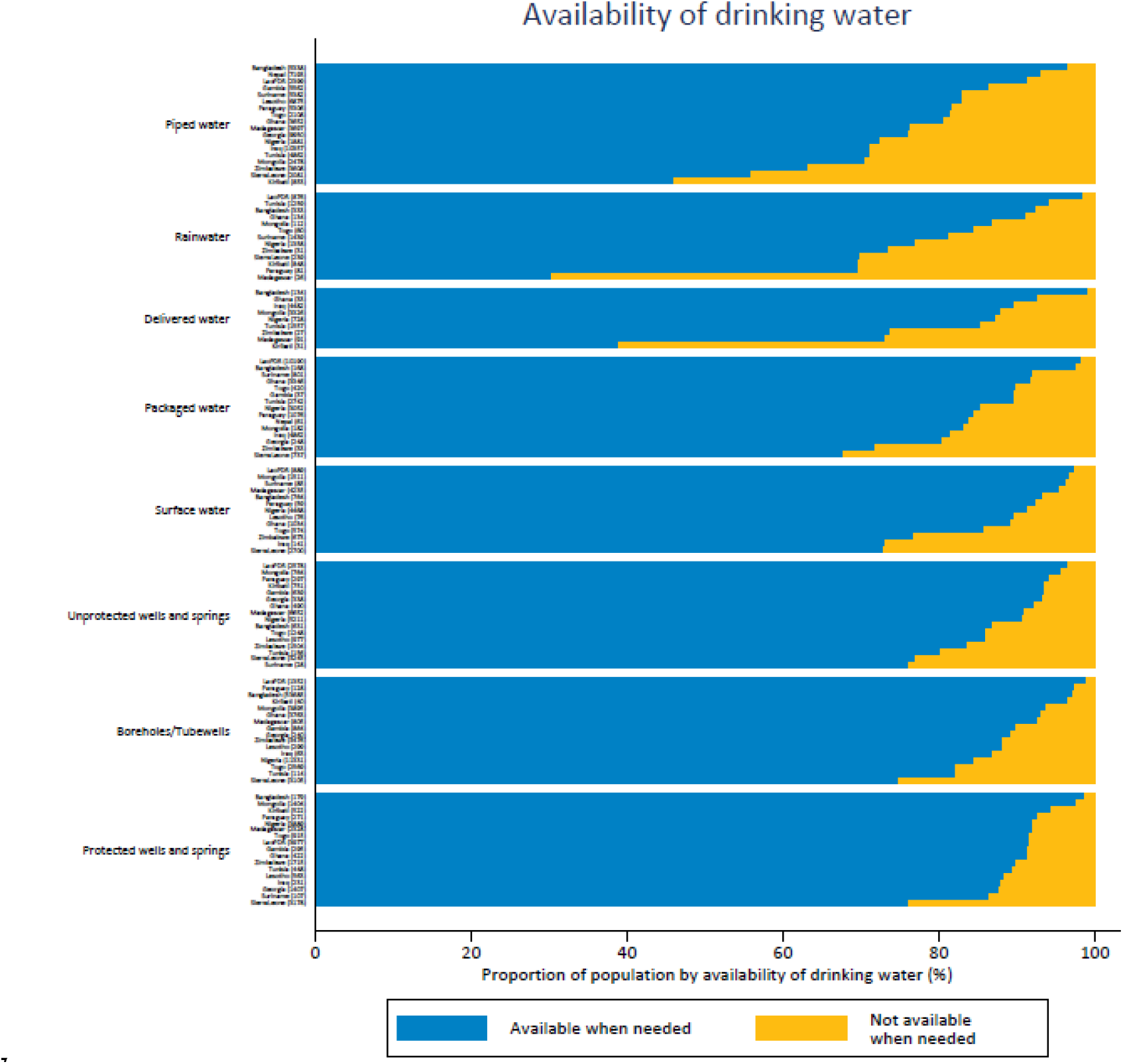
Availability of drinking water, by type of water source. *Note: For source types with at least 25 cases. Water source categories ordered by unweighted mean proportion with available when needed*.

Table S3 lists the main reasons why water was unavailable in the last month. Most households without water in the last month attributed this to water not being available from the source (78.8%) or the source not being accessible (8.5%) and fewer due to water being too expensive (1.5%). A substantial proportion of households without water available reported that there were other reasons, especially in Mongolia (33.0%), Lao PDR (21.8%) and Kiribati (20.5%).

Figure 7 shows the patterns of *E. coli* contamination by wealth quintile. Except in Bangladesh, Kiribati and Sierra Leone, the poorest quintiles are more likely to use sources of drinking water contaminated with *E. coli*. The differences in proportion of the population using sources contaminated with *E. coli* between the richest and poorest quintiles exceeded 50% points in 7 countries. The disparities in water quality at PoU are similar in most of the countries and there is a notable deterioration even in the richest quintile in several countries. With a few exceptions (e.g. Kiribati and Nigeria), absolute disparities between the poorest and richest were generally larger for any contamination than for very high risk drinking water. In contrast, relative inequalities tended to be greater for very high risk drinking water at both PoC and PoU.

**Figure 7:**
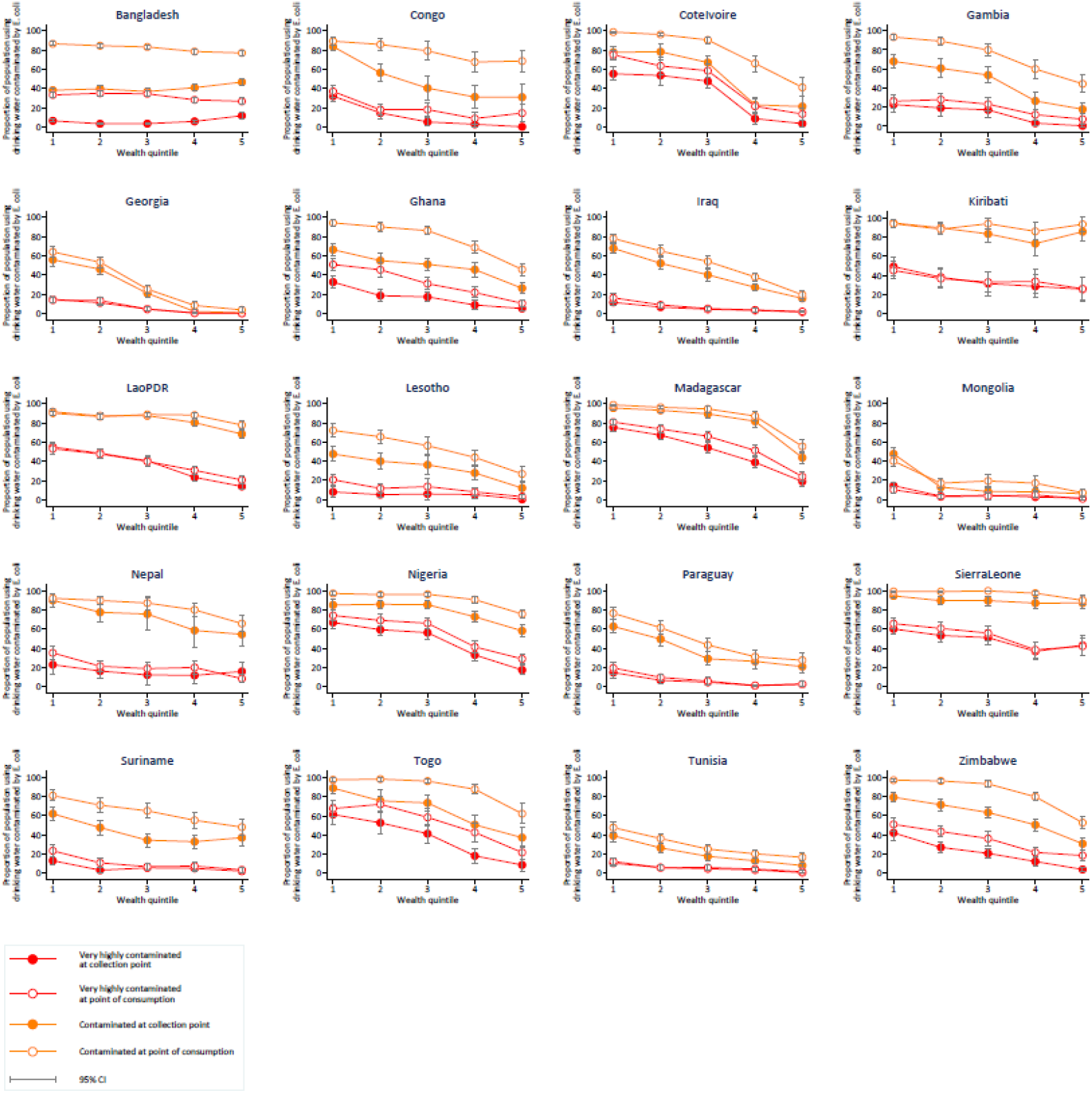
*E. coli* contamination of drinking water at point of collection and point of use, by wealth quintile. *Wealth quintiles from 1 (Poorest) through 5 (Richest)*. Orange lines: At least 1 CFU *E*. coli/100 mL sample; Red lines: Over 100 CFU *E. coli/100* mL.

### Safely managed drinking water services

Table 2 shows national estimates for the proportion of population using improved water sources, as well as the proportion with improved water sources which are accessible, available and free from faecal contamination. In 14 out of 20 countries the quality criterion was the least likely to be met, in the other countries accessibility on premises was the limiting factor. The standard JMP method is to calculate accessibility, availability, and quality of improved drinking water supplies separately in the rural and urban domains, and use the minimum of the three as the safely managed drinking water indicator (WHO/UNICEF 2018a). A national estimate is then produced by weighted average of the rural and urban estimates. This approach allows the JMP to combine data coming from different sources. When information on all of the criteria for safely managed drinking water services are available from a single household survey, they can be combined at the household level to determine where all criteria are met which produces substantially lower estimates.

**Table 2:**
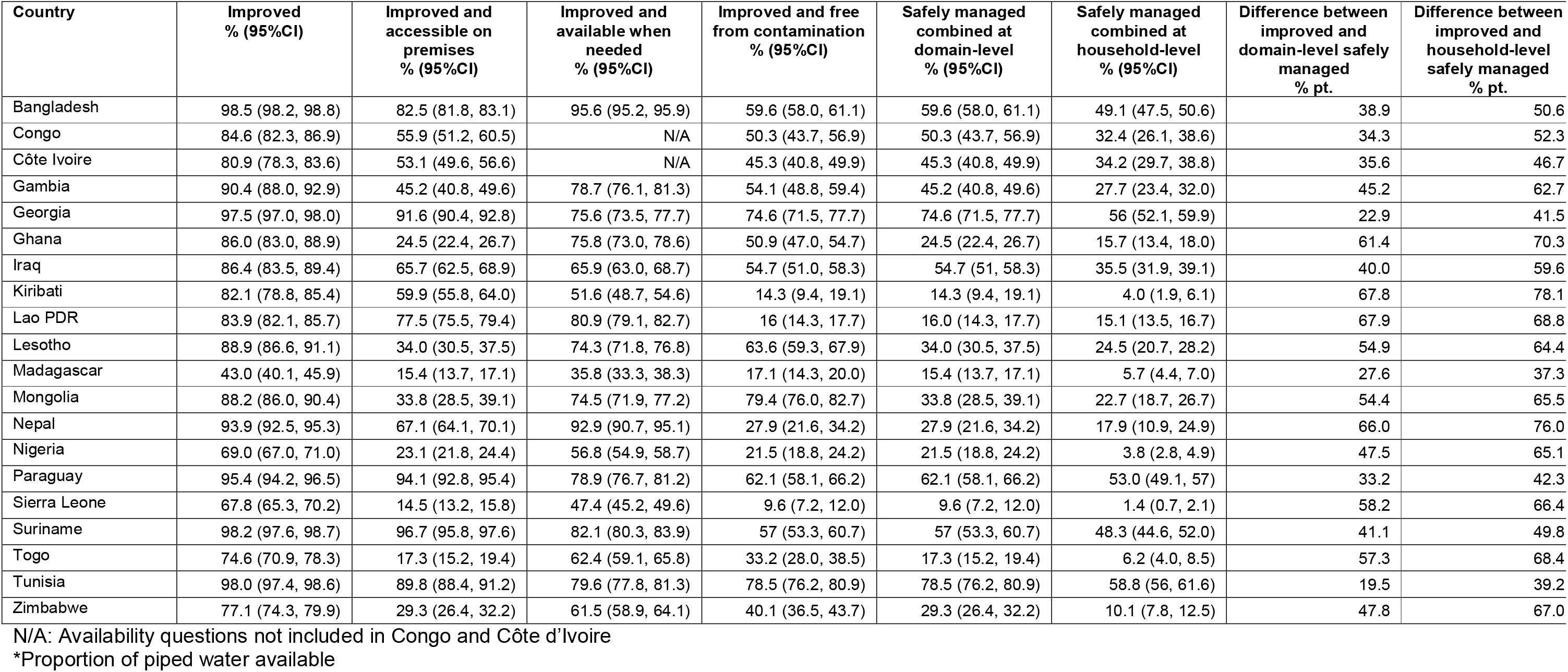
Proportion of population using improved and safely managed drinking water services.

Figure 8 compares the coverage of improved and safely managed services calculated at the domain and household levels. The adjustment for accessibility, availability and quality results in a substantial reduction in all countries, including Bangladesh, Georgia, Paraguay, Suriname and Tunisia where use of improved drinking water sources exceeded 95%. The mean gap between use of improved drinking water and safely managed drinking water services was 58 percentage points calculated at the household level compared with 46 percentage points at the domain level.

**Figure 8.**
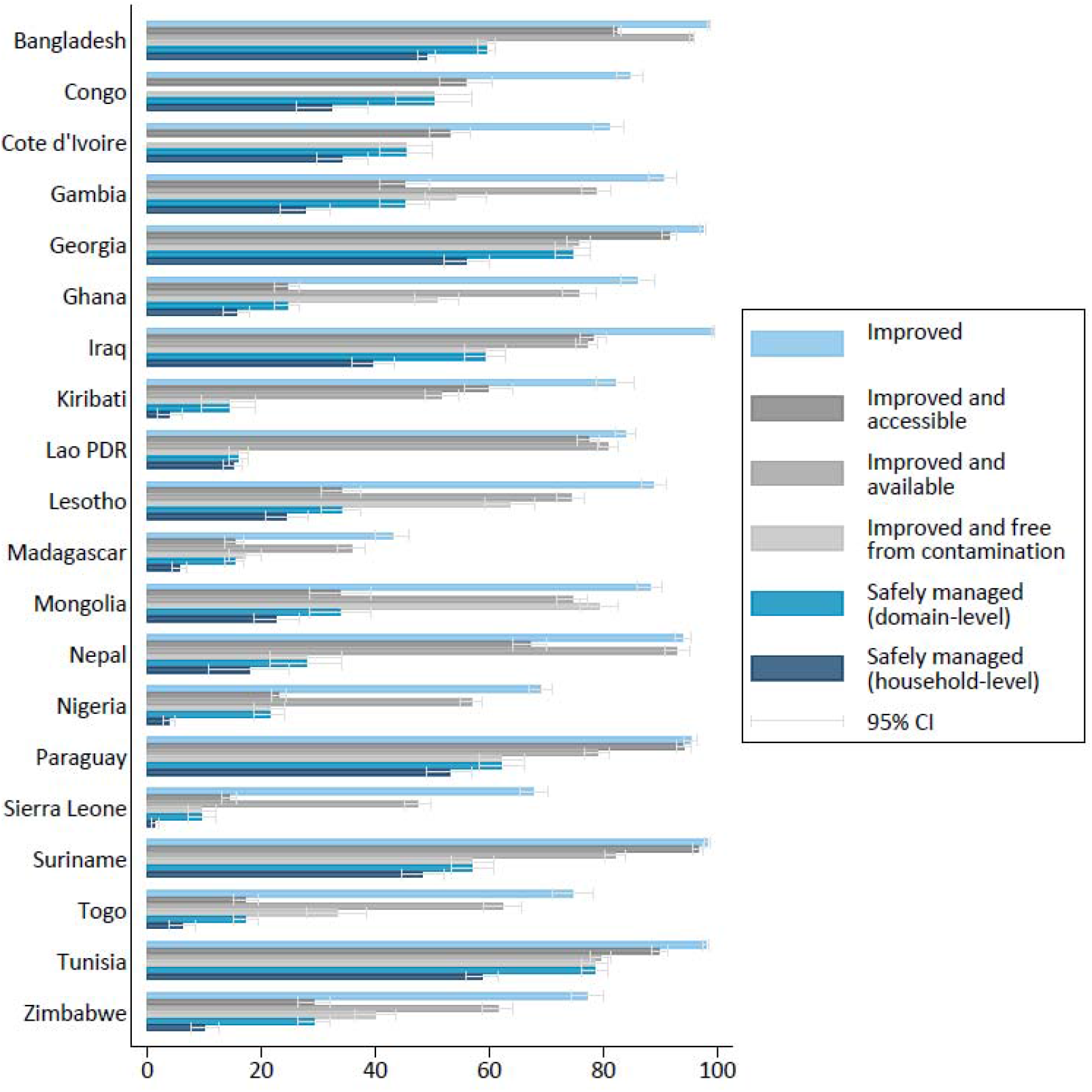
Comparison of improved and safely managed services calculated at domain- and household-level. *Data on water availability in Nepal are for piped water only. No data on availability of drinking water were collected for Congo and Côte d’Ivoire*.

### Risk factors for E. coli contamination

The results of bivariate regression are provided in supplementary materials and we find that all pre-selected variables are significantly associated with faecal contamination at PoC and PoU in at least one country (Tables S9 and S10).

In a first set of multivariate regression models, we examined the association between each of the criteria for safely managed services and faecal contamination at PoC and PoU (Figure 10, Table 3). Figure 10 shows strong and significant impacts of improved water (RR = 0.64 [0.52-0.78]) and accessibility on premises (RR = 0.78 [0.67-0.91]) at PoC. A similar reduction in the risk of contamination at the PoU was seen for water located on premises (RR = 0.75 [0.65-0.86]) but improved sources were less strongly associated at the PoU (RR = 0.85 [0.80-0.92]). RRs for availability of water when needed were all below 1 but were not significant in the pooled estimates at the 5% level. This may reflect the heterogeneity in country-level effect sizes for availability which ranges from Iraq (RR = 0.73 [0.67-0.79]) to Zimbabwe (RR = 1.20 [1.07-1.35]) at PoC. Most RRs were below 1 for individual country models for improved water and accessibility of water on premises (Table 3). Kiribati was the only country with an RRs for PoC contamination that were not significantly below 1 for improved water. RRs for very high levels of contamination are also shown in Figure 11 and demonstrate the protection afforded by improved water sources at PoC (RR = 0.34 [0.24-0.49]) and PoU (RR = 0.58 [0.49-0.67]).

**Table 3:**
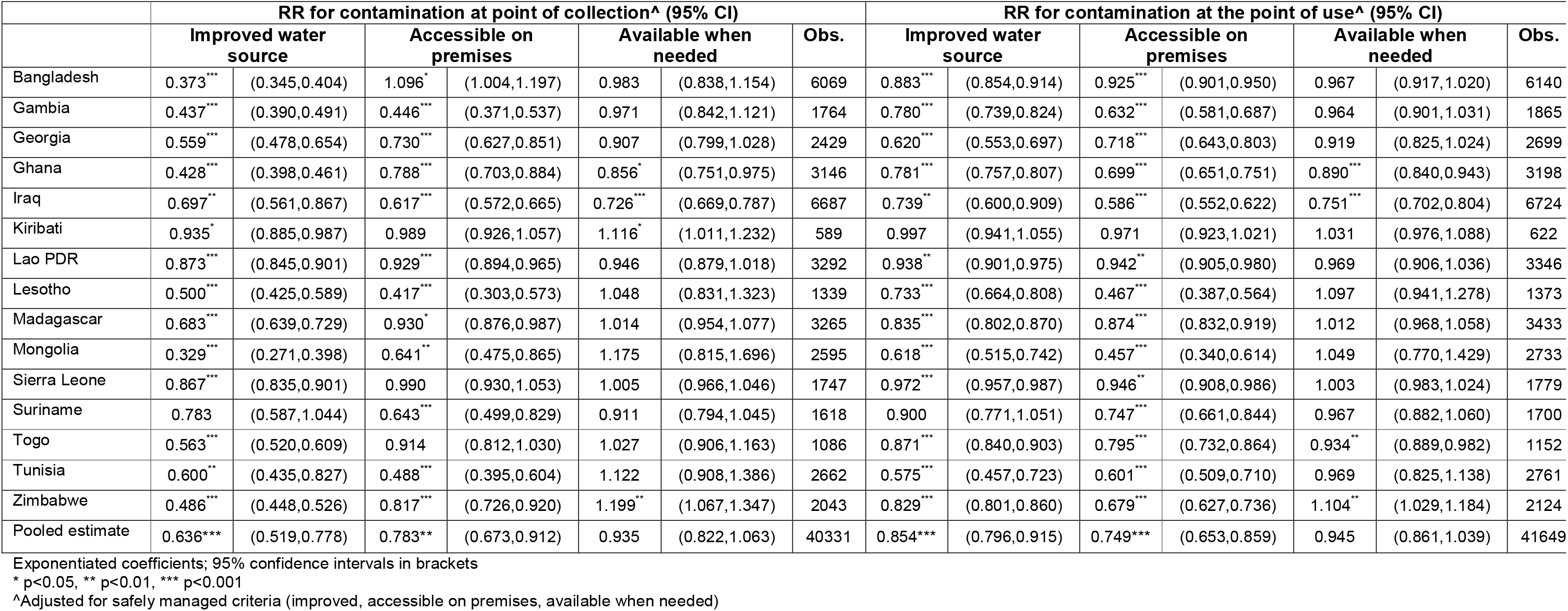
Association between safely managed criteria and faecal contamination.

**Figure 10.**
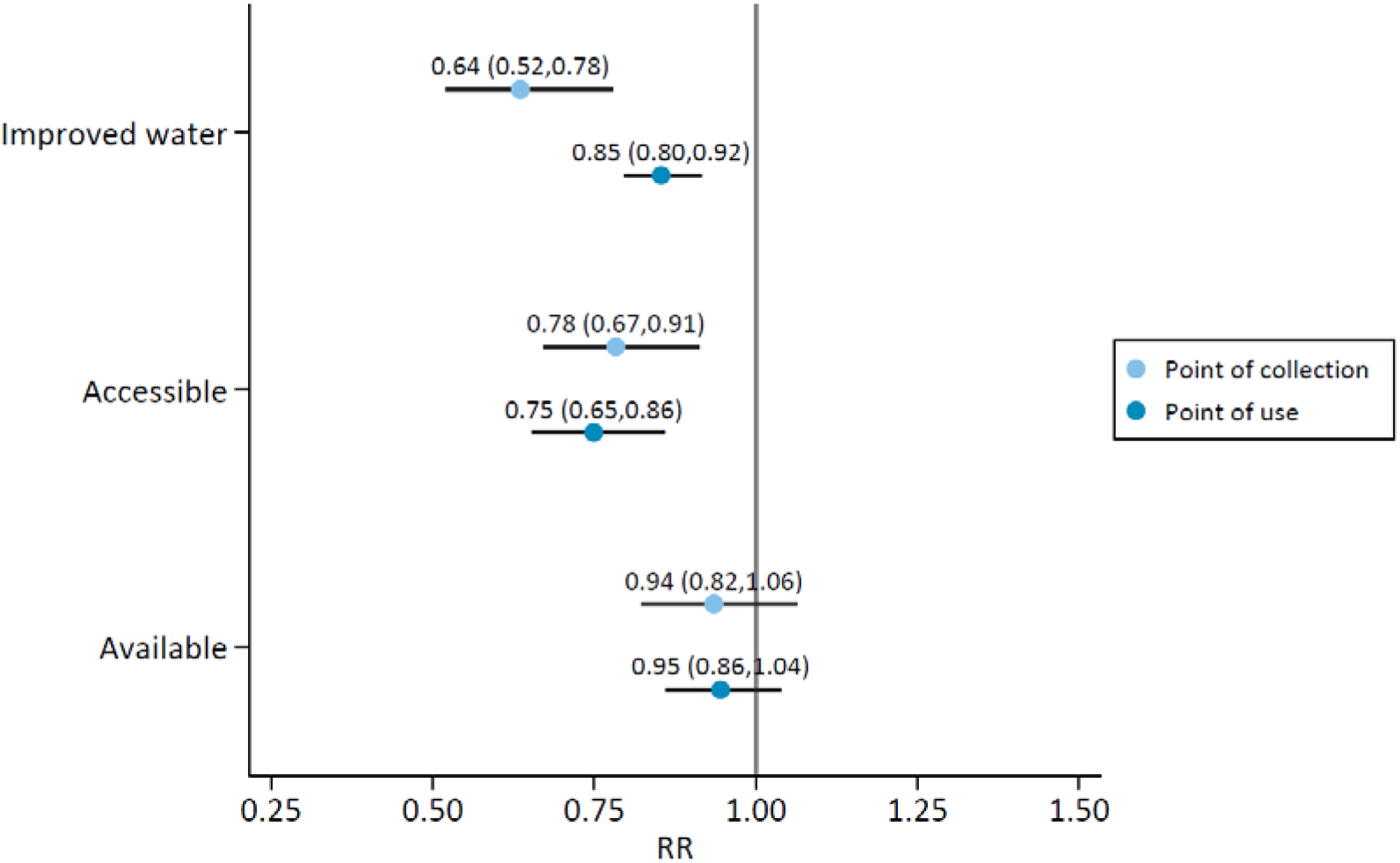
Association between use of improved water sources, and availability and accessibility of improved drinking water sources, and faecal contamination. Includes MICS6 with standard questions on availability of drinking water (n=15). An RR of <1 implies a lower risk of detecting *E. coli*.

**Figure 11.**
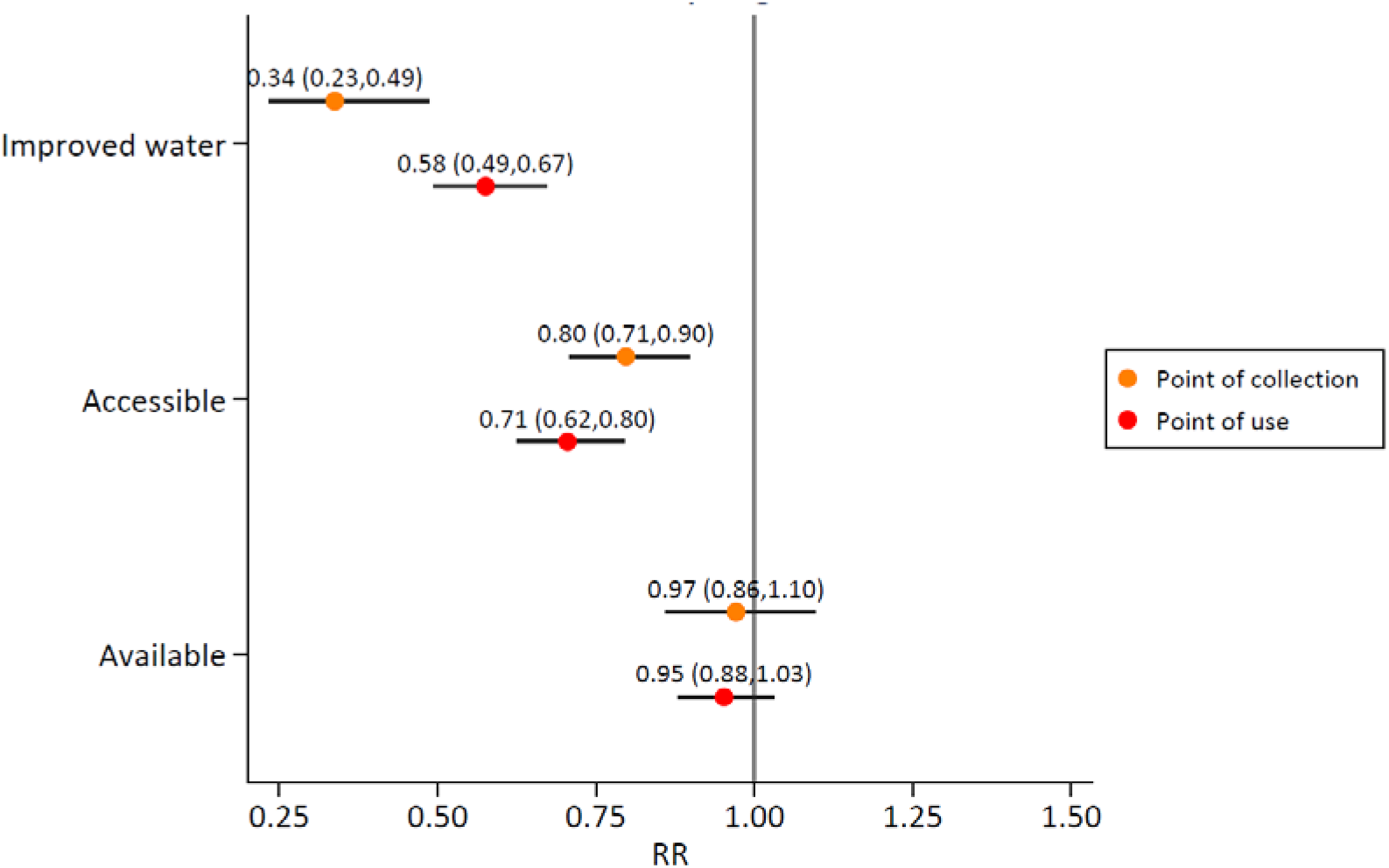
Association between use of improved water sources, and availability and accessibility of improved drinking water sources, and very high risk drinking water. Includes MICS6 with standard questions on availability of drinking water (n=15). An RR of <1 implies a lower risk of > 100 *E. coli* per 100 ml.

In a second set of multivariate regression models we included all pre-selected risk factors for contamination. The RRs for contamination and very high risk drinking water vary considerably between countries with each risk factor significantly associated with faecal contamination in at least one country (Figures S12-S26). Figure 12 and 13 shows the pooled results across the 15 MICS6 and demonstrates the strong impact of water supply type and wealth in these models and confirms the risk of contamination is considerably lower for some types of improved source than others. Compared to unimproved water sources, pooled estimates suggest that piped water 0.58 (0.44,0.76) (RR = 0.60 [0.51-0.71]), boreholes/tubewells 0.61 (0.51,0.73) (RR = 0.59 [0.47-0.74]) reduced the risk of contamination at PoC whereas protected dug wells and springs 1.00 (0.93,1.08) (RR = 1.0 [0.94-1.10]) and rainwater 1.10 (0.86,1.42) (RR = 1.1 [0.88-1.30]) did not. At PoU only piped water was found to be associated with a lower risk of contamination 0.87 (0.81,0.93) (RR = 0.86 [0.80-0.92]). Water accessible on premises was associated with decreased risk of contamination at PoU 0.92 (0.88,0.96) (RR= 0.92 [0.88-0.96]). All improved drinking water source types offered protection against very high risk drinking water at PoC and PoU, with the exception of rainwater and delivered water which were not significant at PoU.

Compared to the poorest quintile, the risk of very high risk contamination at PoU was lower in all other quintiles, with a clear trend with increasing wealth. Animal ownership (RR = 1.08 [1.03-1.14]) and rural residence (RR = 1.11 [1.03-1.19]) were found to increase the risk of contamination at the PoC. Community (>75%) improved sanitation coverage 0.95 (0.91,0.98) (RR = 0.95 [0.92-0.99]) but not household sanitation coverage 1.03 (0.96,1.10) (RR = 1.02 [0.99-1.06] was associated with lower risk of contamination at PoU. Household water treatment was not significantly associated with lower risk of contamination at PoU (RR = 0.90 [0.79-1.03]).

**Figure 12.**
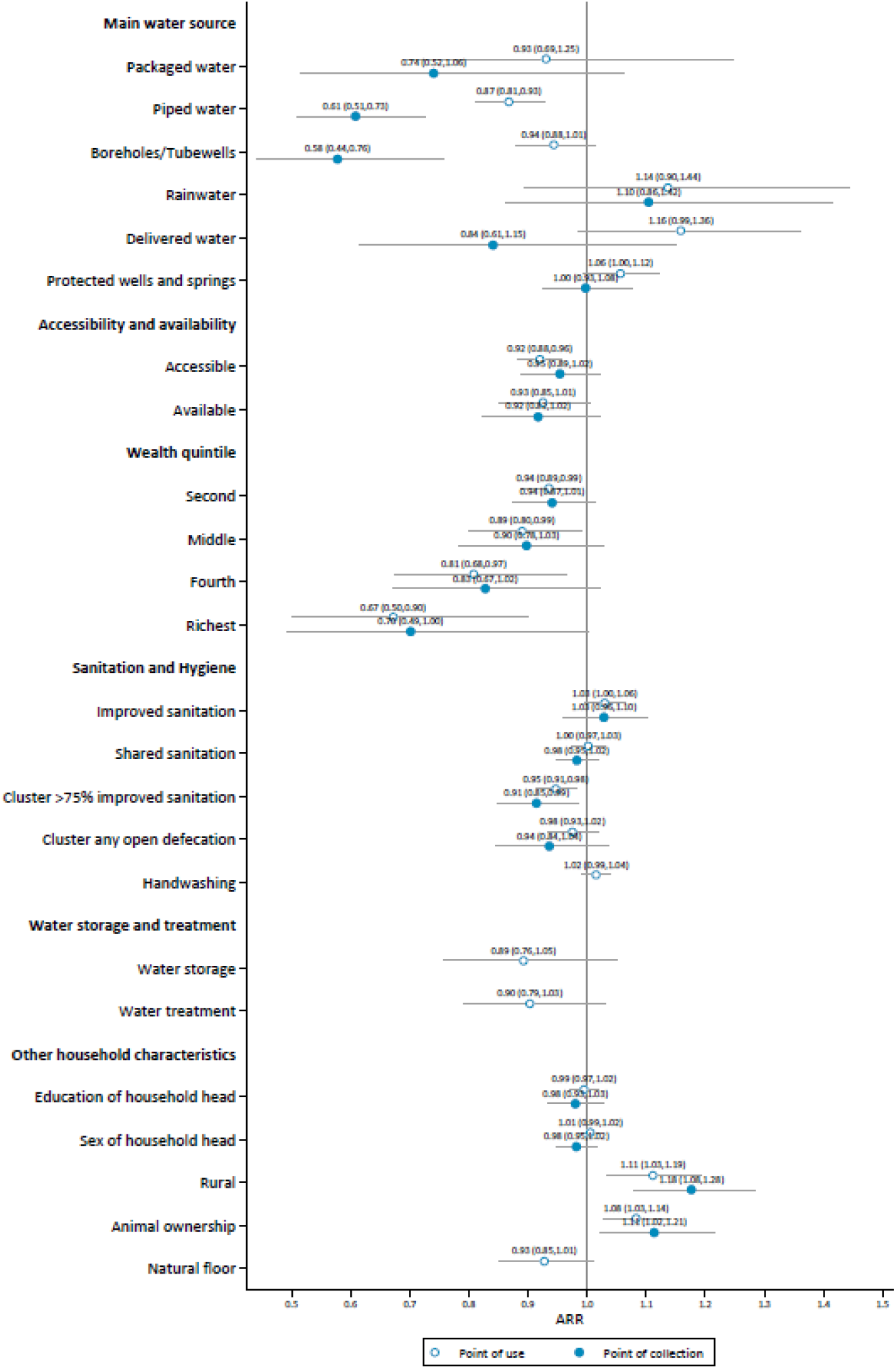
Adjusted relative risks for faecal contamination of drinking water at point of collection and point of use. Includes MICS6 with standard questions on availability of drinking water (n=15). An RR of <1 implies a lower risk of detecting *E. coli*.

**Figure 13.**
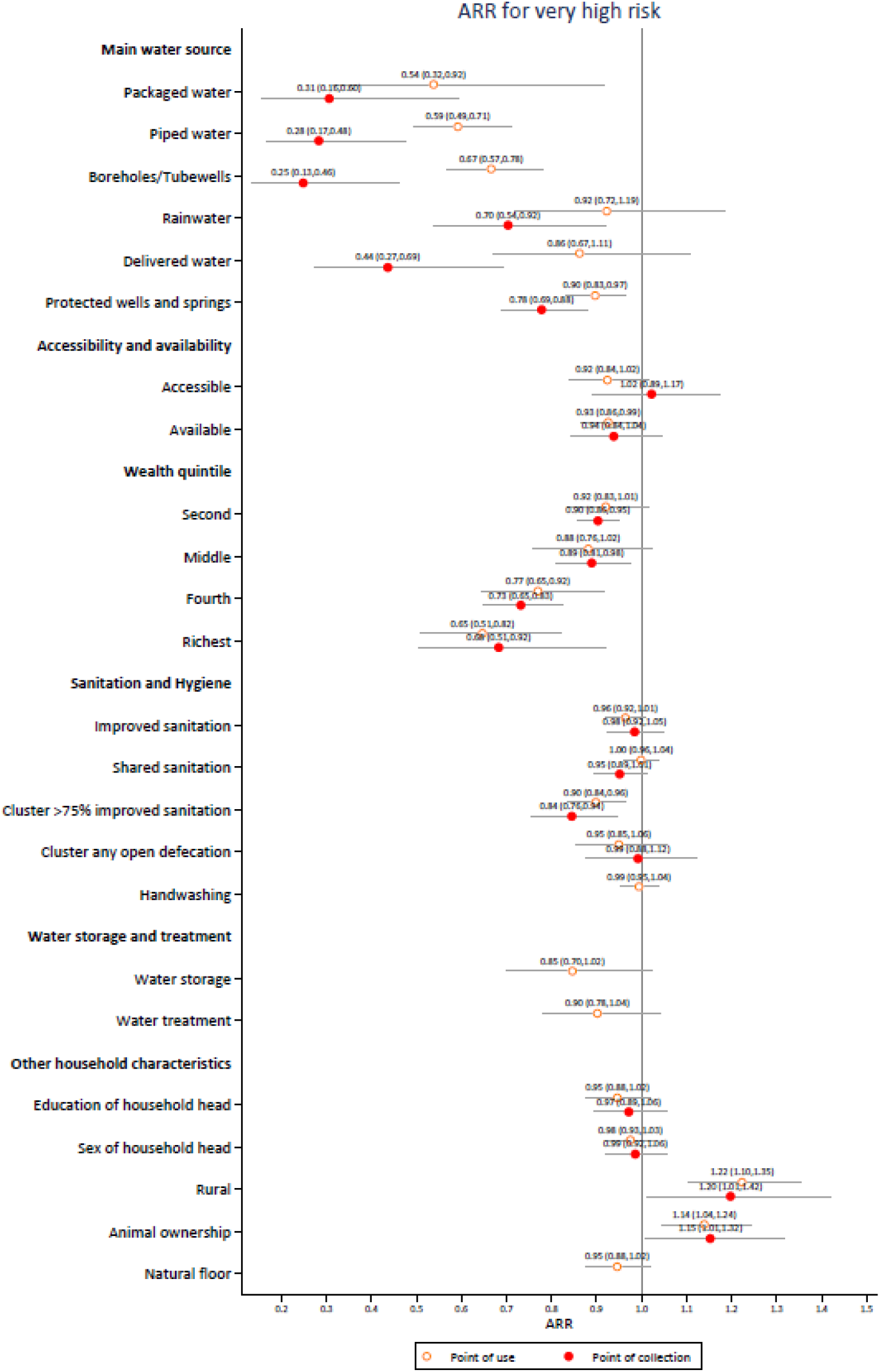
Adjusted relative risks for very high risk drinking water at point of collection and point of use. Includes MICS6 with standard questions on availability of drinking water (n=15). An RR of <1 implies a lower risk of > 100 *E. coli* per 100 ml.

In sensitivity analyses we found excluding WASH variables from the wealth index marginally increased the RRs for water supply types and decreased the strength of the association with wealth (Figure S27) however use of the source of the water quality sample rather than a household’s main source had a negligible impact on RRs for water source types (Figure S28).

## Discussion

The new SDG indicator for drinking water "safely managed services” reflects consensus on the need to go beyond monitoring the types of water sources used by households and to monitor the level of service received. Integration of water testing in multi-topic household surveys has demonstrated the feasibility of this approach for collecting representative data on drinking water service levels and the ability to link them with information on the socio-economic characteristics of the population. Integrating water testing into existing surveys provides a cost-effective approach compared with running a dedicated water quality survey.

### Water quality and risk factors for contamination

The findings in the countries included in the analysis show that large proportions of the population are exposed to faecal contamination through their drinking water and many people are exposed to very high levels of contamination.

In line with a systematic review of the quality of water from different sources (Bain et al. 2014b), we find that piped water, packaged water and boreholes are generally providing higher quality water than unimproved sources and they offer substantial protection against very high risk drinking water. Rainwater harvesting, delivered water, and protected wells and springs often do not provide drinking water that is less likely to be contaminated than unimproved sources. Piped water was the only source type significantly associated with water free from contamination at PoU in the adjusted models.

By combining data on water quality with information on the socio-economic status of households, it was possible to explore patterns by wealth quintile in greater detail that prior studies (Yang et al. 2013). We find pronounced differences between rich and poor in almost all countries. Differences were smaller in Bangladesh where the richest were found to be more likely to use water from a contaminated source. This reflects the high degree of contamination of piped water supplies in Bangladesh disproportionately used by the wealthy, compared to the less contaminated boreholes used by most of the population.

The surveys also confirm that there is often substantial deterioration in quality between PoC and the point where households consume drinking water (Shields et al. 2015; Wright et al. 2004). In 25% to 40% of households the water quality deteriorated by at least one risk class, compared with improvements in only 8% to 20%. In many countries appropriate household water treatment practices were relatively uncommon and most households store drinking water at home. Our analysis suggests that household water treatment usually does not provide substantial protection for households using contaminated sources (with the notable exception of boiling in Mongolia). These findings are likely to be impacted by a lack of correct and consistent use of water treatment practices and reporting bias, which has been documented in other countries (Rosa et al. 2016; Rosa and Clasen 2017), and call into question the use of household water treatment in burden of disease estimates by WHO and IHME (Pruss-Ustun et al. 2019; Stanaway et al. 2018). In contrast, we find that animal ownership (Wardrop et al. 2018), rural residence and community sanitation coverage are significantly associated with contamination at PoC and PoU in the adjusted regression models.

The risk factors for contamination vary considerably between countries and this heterogeneity is reflected in the range of effect sizes in the individual country models. Further work is required to understand the relationship between household and community level risk factors and the extent to which these can be mitigated through interventions to improve infrastructure and change behaviours.

### Monitoring safely managed services

In addition to integrating the water quality module, questions that address the availability and accessibility of drinking water are required to monitor safely managed services. Questions on the location and/or time to collect drinking water have been included since the second round of MICS in 1999-2003 and form part of the JMP’s updated guidance on monitoring WASH in household surveys (WHO/UNICEF 2018b). New questions to assess the availability of drinking water services were asked in 17 out of 20 countries to establish the population lacking sufficient drinking water when needed. In these countries the proportion of the population drinking water that was not accessible on premises, or which was contaminated with *E. coli*, was generally much higher than the proportion reporting lacing sufficient water in the last month.

Multivariable regression analysis exploring the association between drinking water sources meeting the criteria for safely managed services provides further evidence that improved drinking water services are more likely to be free of contamination and underscores the importance of accessibility on premises for water quality, especially at the PoU. However, the models suggest that improved sources meeting the criterion for availability are not necessarily more likely to be free from contamination. Furthermore, the relationships between the individual criteria differ between and within countries. This suggests that it is important to separately monitor these new criteria and that there may be tradeoffs in seeking to improve quality, availability or accessibility.

The integration of water quality testing in household surveys should complement, not compete with, ongoing efforts to strengthen surveillance of water quality by regulatory authorities. In many low and middle-income countries, water quality data from regulatory authorities are limited, especially for rural areas. Nationally representative data from household surveys can provide a cost-effective means of filling these data gaps in the short term and draw attention to service quality in the absence of regulation. Furthermore, the responsibility of the service provider usually ends at the household connection or public tap, as a result regulators rarely collect samples from within the home.

### Recommendations for water testing in future household surveys

The water quality module has been integrated in an increasing number of MICS and other national and sub-national surveys with support from the JMP team including national household surveys in Ghana, Democratic People’s Republic of Korea, Ethiopia, Ecuador and Lebanon (WHO/UNICEF 2019a). There have also been sub-national pilots in the Demographic and Health Survey in Peru (Wang et al. 2017), SUSENAS in Indonesia (Cronin et al. 2017) and Living Conditions Survey in Afghanistan (Central Statistical Organization of Afghanistan 2017). This illustrates the growing demand for understanding the quality of services and the willingness of national authorities to shine a light on this important but politically sensitive issue.

Several key lessons from these experiences taking water testing to scale could inform the design and implementation of future surveys. In particular, testing drinking water in a sub-sample of households for a limited number of priority parameters is critical to the cost effectiveness and practicability of this approach. The design effect will vary depending on the number of samples tested but also the context, parameter and survey stratification. Analysis of these early surveys suggests that conducting water quality testing in approximately four households per cluster provides a good balance between increasing statistical power and minimizing the design effect. Strong technical support is often available from national authorities and water quality experts who are familiar with membrane filtration in a laboratory setting. Involvement of regulatory authorities is strongly recommended given their mandate for oversight of water service provision. Quality control measures are important during fieldwork and in order to build confidence in the results – blank tests are especially valuable to confirm that detection of *E. coli* is not the result of poor hygiene by field teams. Close attention is needed for the training which, depending on the number of teams and facilitators, should last 3-5 days and include sufficient practice (preferably at least 15 tests) for each step in the process.

Due to logistical considerations and concerns related to misinterpretation of test results by households, water quality data were not shared with individual households but were made available in aggregate form in the MICS final reports and in anonymized form in the datasets used in this analysis. In a few countries leaflets explaining contamination risks and methods for safe water treatment and storage were provided to households selected for the water quality module. Further work is needed to determine how best to share information on water quality with households and their communities in order to inform decisions on data sharing by implementing agencies (Khan et al. 2017).

### Water testing equipment and procedures

There are several water quality tests that could potentially be integrated in household surveys (Bain et al. 2012). In the approach used in MICS an adapted membrane filtration process has been used, yielding reliable quantitative results (Brown et al. 2020). The costs are approximately $2.5 per test using the new *"E. coli* only” Compact Dry plates and $1500 per team for hardware (mainly the filtration equipment) but these are expected to reduce over time.

The use of enzymatic substrates offers specific detection of *E. coli* and permits the use of non-standard incubation temperatures in contrast to the often-used thermotolerant coliform tests which require incubation at circa 44⍰C to ensure specificity (Matthews and Tung 2014). Although incubation belts have proven to be a pragmatic approach, further research and development on incubation technologies is merited since belts need to be worn overnight and are considered inappropriate in some parts of the world (e.g. northern Nigeria, Afghanistan). Ambient temperature and phase change incubators are both potential solutions (Brown et al. 2011) and CompactDry™ plates have been shown to perform favourably when compared to reference methods when incubated at ambient temperature in Bangalore, India (Brown et al. 2020).

Future surveys could consider the use of presence/absence testing as used in Ecuador ENEMDU (Moreno et al. 2020)(where contamination is expected to be relatively rare) or combined with a 1 mL plate (to detect high levels). Alternatively, the reuse of manifolds in multiple surveys or the development of a lower cost filtration apparatus could significantly lower costs for surveys with larger numbers of teams such as Nigeria. A low-cost manifold (<US$100) has successfully been piloted in the Afghanistan Living Conditions survey (Central Statistical Organization of Afghanistan 2017) and provincial MICS in Pakistan (Pakistan Punjab Bureau of Statistics 2018). In the longer term it is hoped that novel, rapid tests will replace the culture-based approaches that dominate the water quality testing market (Rompre et al. 2002; UNICEF 2017). Such tests have the potential to greatly reduce the cost and complexity of water testing in household surveys.

### Limitations

Integrating water quality testing in nationally representative household surveys provides a single measurement of water quality, often during a season when weather is favourable for fieldwork, and is not a substitute for routine monitoring by the responsible authorities in each country. As a snapshot of quality it will likely underestimate exposure to faecal indicator bacteria ‘at all times’. Furthermore, *E. coli* as an indicator of faecal contamination has known limitations (Gleeson and Grey 1996) including greater sensitivity to chlorine than pathogens such as *Cryptosporidium* (WHO 2017).

To match published reports, our main analysis of inequalities by wealth did not exclude water, sanitation and hygiene facilities in the definition of the asset index (Martel 2017) but we conducted a sensitivity analysis to explore this. The sample size, especially in Nepal and Kiribati, limits the precision of estimates for subpopulations and inference from the regression models. Different concepts of availability were evaluated in the MICS5 round, limiting comparability with data collected during the MICS6 round and highlighting the importance of standardized approaches to data collection. The exploratory regression analysis examined the presence or absence of *E. coli* and exposure to very high risk drinking water (>100 *E. coli* per 100 mL) at PoC and PoU and future studies could examine changes in risk levels between these two sampling locations in more detail. There are many potential risk factors for water quality that were not considered in the regression analysis either because they are not collected in MICS (e.g. metrological data, sanitary risk assessments) or because the number of missing values is large (e.g. child faeces disposal practices). Furthermore, the cross-sectional nature of the data may understate the importance of risk factors that may be important during the wet season. Regression estimates were not weighted to reflect population sizes and the contribution of each survey to the pooled estimates reflects the varying number of samples taken in each country.

## Data Availability

All data are available from the UNICEF Multiple Indicator Cluster Survey website.

http://mics.unicef.org/surveys

## Acknowledgements

We are grateful to the implementing agencies who integrated water testing in national household surveys. We thank the entire MICS team for the excellent collaboration in the development of the new water quality module as well as the JMP water quality trainers providing technical support to countries implementing the water quality module. The following international consultants supported water quality testing in MICS: Abdus Saboor, Andrew Shantz, Caetano Dorea, Elisabeth Lictevout, Lars Osterwalder, Loay Hidmi, Mamadou Djerma and Peter van Maanen. Yadigar Coskun provided data processing support on the custom wealth indices. Jennyfer Wolf, Matt Freeman, Andrew Mertens and Amy MacDougall provided guidance on multilevel regression modeling. Jennifer de France and Angella Reinhold kindly reviewed a draft of the study.

## Funding

Funding for the integration of water testing in household surveys was provided by the Netherlands Directorate-General of International Cooperation (DGIS), the UK Department for International Development (DFID) and the United States Agency for International Development (USAID).

## Competing Interests

RB TS RJ work for the WHO/UNICEF Joint Monitoring Programme. AH and SK work for the UNICEF Multiple Indicator Cluster Survey Programme. Authors’ opinions do not reflect position of WHO or UNICEF.

## Notes

### Author Declarations

Office of Research Compliance & Outreach, Baruch College determined that this study was exempt from IRB/HRPP review.

